# Use of assistive technology to assess distal motor function in subjects with neuromuscular disease

**DOI:** 10.1101/2024.05.19.24307604

**Authors:** Dominique Vincent-Genod, Sylvain Roche, Aurélie Barrière, Capucine de Lattre, Marie Tinat, Eelke Venema, Emmeline Lagrange, Adriana Gomes Lisboa de Souza, Guillaume Thomann, Justine Coton, Vincent Gautheron, Léonard Féasson, Pascal Rippert, Carole Vuillerot

## Abstract

**Purpose:** Among the 32 items of the Motor Function Measure scale, 3 concern the assessment of hand function on a paper-based support. Their characteristics make it possible to envisage the use of a tablet instead of the original paper-based support for their completion. This would then make it possible to automate the score to reduce intra- and inter-individual variability.

The main objective of the present study was to validate the digital completion of items 18, 19, and 22 by measuring the agreement of the scores obtained using a digital tablet with those obtained using the original paper-based support in children and adults with various neuromuscular diseases (NMD). The secondary objective is to calibrate an algorithm for the automatic items scoring.

**Design:** Prospective, multicentre, non-interventional study.

**Methods:** Ninety-eight subjects aged 5 to 60 years with a confirmed NMD completed MFM items 18, 19, and 22 both on a paper support and a digital tablet.

**Results:** The median age of included subjects was 16.2 years. Agreement between scores as assessed using the weighted Kappa coefficient was almost perfect for the scores of items 18 and 22 (*K*=0.93, and 0.95, respectively) and substantial for item 19 (*K*=0.70). In all cases of disagreement, the difference was of 1 point. The most frequent disagreement concerned item 19; mainly in the direction of a scoring of 1 point less on the tablet. An automatic analysis algorithm was tested on 82 recordings to suggest improvements.

**Conclusion:** The switch from original paper-based support to the tablet results in minimal and acceptable differences, and maintains a valid and reproducible measure of the 3 items. The developed algorithm for automatic scoring appears feasible with the perspective to include them in a digital application that will make it easier to monitor patients.

## Introduction

In subjects affected by a neuromuscular disease (NMD), muscle damages progressively affect motor function leading to limitations in the activities of daily living. The assessment of motor function is crucial to measure the impact of the disease on functional limitations and participation restrictions in order to propose adequate management of people living with a NMD [1–3]. In subjects with the most severe phenotypes, assessment is focused on upper limb distal motor function as the distal muscles groups of the upper extremities are those best preserved for the longest. Studies have found that adult subjects with Duchenne muscular dystrophy are still able to perform important functional activities despite having limited distal motor function. Over time, they tend to lose their functional abilities, although their muscle strength decreases only slightly [4,5]. Upper limb function measurement is also of value for certain NMD where upper limb distal motor function is primarily impaired, such as the distal myopathies and peripheral neuropathies, for routine monitoring of such patients but also clinical trials [6]. In addition, upper extremity function is determined not only by weakness, but also by prominent and progressive joint contractures or sometimes prominent distal laxity underscoring the need for valid hand motor assessments applicable in various NMD [7]. The Motor Function Measure (MFM) is a 32-items scale for assessing motor function of subjects with a NMD that is frequently used in clinical trials and clinical follow-up. Validation studies have confirmed its value, its ease of use, validity, reproducibility, and responsiveness [8,10]. Despite the many advantages and increasing use of functional assessment scale, there is still a variation in ratings between examiners and by the same examiner, linked to the subjectivity and intrinsic qualities of the assessor, a limitation found in all hetero functional assessments.

The characteristics of 3 items focused on upper limb distal motor function make it possible to envisage using a digital tablet instead of the original paper support to administer them. Digitising the 3 items paved the way to the automatization of the item scoring, diminishing the therapist’s influence on item rating levels.

In the present study we present a validation study of the digital completion of items 18, 19, and 22 by measuring the agreement of the scores obtained using a digital tablet (T-score) with those obtained using the original paper-based support (P-score) in children and adults with various NMD, as well as assessing performances of automatic items scoring algorithms.

## Methods

### Study design and ethic

This prospective, multicentre, non-interventional study conducted in 5 neuromuscular disease reference centres located in 3 university hospitals; 2 paediatric and 3 adult centres. Patients aged 5 to 60 years with a confirmed NMD and scheduled to be evaluated using the MFM were invited to participate during a clinical follow-up visit. They were provided with written information describing the study and, according to legislation in place at the time of the study, patients (or parents/legal guardian) who gave oral consent to participate were included between January to December 2018. Ethics approval was obtained from the regional review board (*Comité éthique de Protection des Personnes du Sud-Ouest et Outre-Mer de France*), the study was registered on ClinicalTrials.gov (ID: NCT03465358).

### The MFM scale

The MFM is a functional rating scale available in 2 versions; one composed of 32 items dedicated to subjects with a NMD aged 6 to 60 years (MFM-32)[8,10], the other composed of 20 items of the MFM-32 validated for children aged from 2 to 7 years of age (MFM-20)[11]. In both versions, items are grouped into 3 domains: D1, standing and transfers; D2, axial and proximal motor function; and D3, distal motor function. The scoring of items uses a 4-point Likert scale based on the individual’s maximal abilities. The generic scoring is 0= cannot initiate the exercise or maintain the starting position; 1= performs the task partially; 2= performs the movement incompletely, or completely but imperfectly (compensatory movements, slowness …); and 3= performs the task fully and ‘normally’ (the detailed rating procedure, is described in the MFM user’s manual available from http://www.mfm-nmd.org). The total score, as well as the D1, D2, and D3 sub-scores, are expressed as a proportion (%) of the maximum possible score; the lower the score, the more severe the impairment.

MFM items 18, 19, and 22 are items performed originally on a paper-based support, with the subject sitting on a chair or in a wheelchair in front of a table.

The task to be perform for item 18 is to go round the edge of a CD, with one finger placed in the centre of the CD at the beginning of the item. The CD is glued to a piece of cardboard (Fig 1A). The scores are as follows: 0, the subject cannot go round the small circle of the CD (diameter 3.5 cm) with a finger; score 1, he goes round the small circle of the CD with a finger; score 2, he goes round the edge of the CD with compensatory movements or difficulty, he may stop one or more times and/or may change finger during the task, and compensatory pf the trunk are allowed; score 3, he goes round the edge of the CD with the same finger without hand support on the table and without compensatory movements.

**Fig 1.**
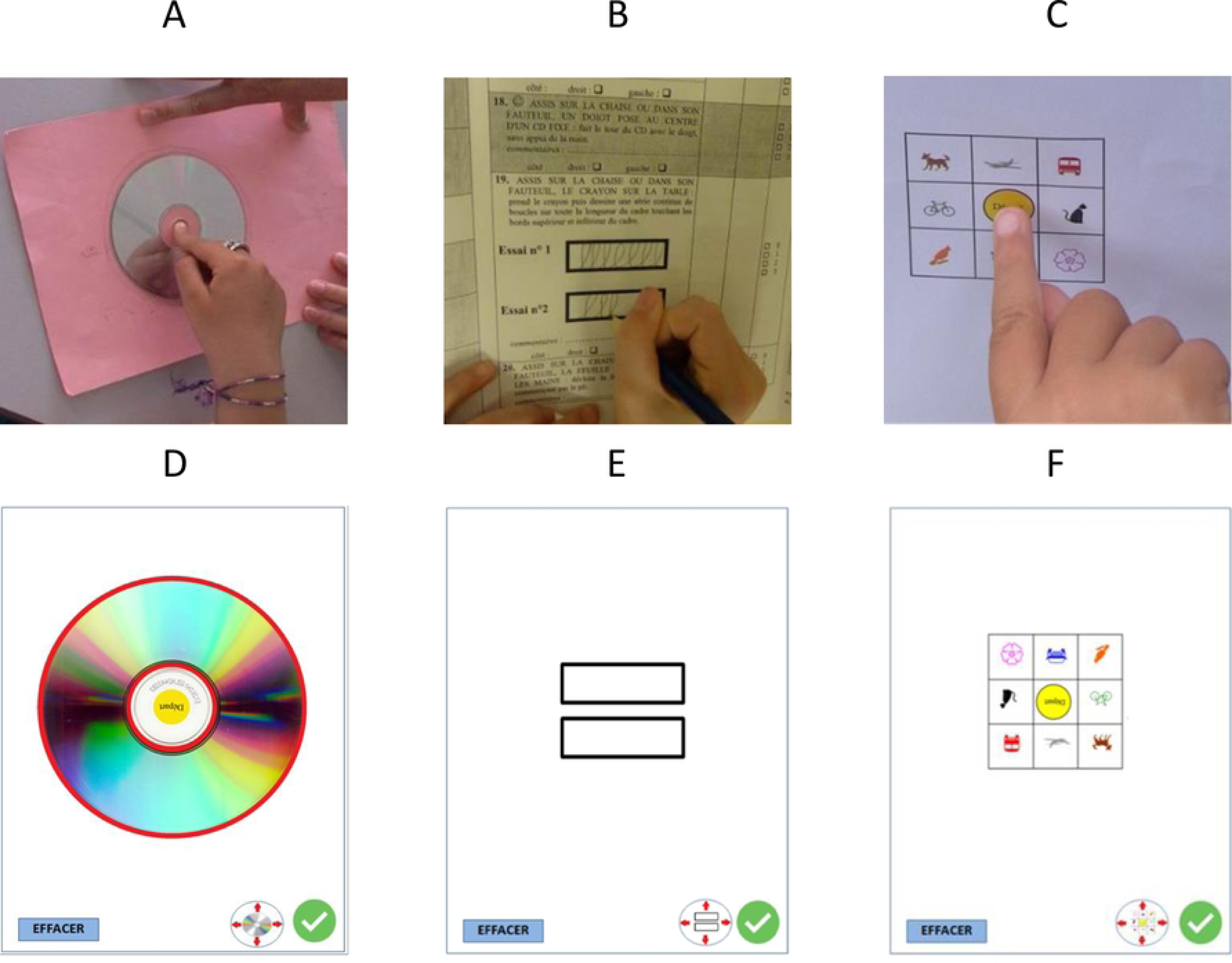
Original paper-based support for item 18 (A), 19 (B) and 22 (C) and screenshots of the digital interfaces of the item 18 (D), 19 (E) and 22 (F)

The task to perform for item 19 is to pick up a pencil and draw loops inside a frame (Fig 1B). The frame is a rectangle measuring 1 cm high and 4 cm long, with a line width of 0.1 cm. The scores are as follows: 0, the subject cannot pick up the pencil or cannot make a written mark; score 1, he cannot draw one loop inside the frame touching the top and bottom of the frame; score 2, he draws at least one loop inside the frame, but cannot draw a continuous series of loops in the frame touching the top and the bottom lines of the frame; score 3, he draws a continuous series of loops over the full length of the frame without stopping.

The task to perform for item 22 is to place a finger on drawings in a diagram from an initial touch on the centre of the diagram (Fig 1C). The scores are as follows: 0, the subject cannot raise the finger, nor slide it onto a drawing; score1, he cannot raise the finger to place it on a drawing, but can slide it on at least one drawing; score 2, he raises the finger and places it imprecisely on 1 to 8 drawings on the diagram; score 3, he raises the finger and places it successively on the 8 drawings of the diagram without touching the lines. For all scoring levels, the hand and/or the other fingers may give support.

Items 18 and 22 are in both the MFM-32 and MFM-20; item 19 is only in MFM-32.

### Choice of the tablet

The ZenPad 3S 10 Android tablet associated with the Z Stylus (Asus, New Taipei, Taiwan) was used in the present study. The tablet has a 9.7-inch screen (sufficient to contain the dimensions of a CD), it is 5.8mm thick, has a resolution of 1536 X 2048 pixels, and a multi-touch capacity allowing the tracing of 5 fingers simultaneously; the Z Stylus has a peak of 1.2 mm, and weighs 11.58 g.

### TabMe2 software

In order to record the subject’s traces left on the ZenPad tablet, a software (TabMe2) was developed by the G-SCOP laboratory (Grenoble, France). It allows recording the X and Y coordinates over time of fingers or stylus on a tablet and includes a module to display the recorded tracks retrospectively. The digital interfaces have been developed respecting the size and symbols of the “materiel” usually used in the MFM (Fig 1). The figures representing the CD, the rectangle and the diagram can be moved and positioned manually on the screen according to the subject’s preferred choice. The ‘EFFACER’ button can be used to reset the plot and perform the evaluation any number of times.

Mathematical models and decision-making algorithms were applied on the recorded traces (see S1 File.) to give automatic scorings. The automatic scoring procedures were developed during the course of the study and could only be applied to the last patients included.

### Study procedure

Subjects aged 6 years or more completed the MFM-32 in the usual way, and items 18, 19 and 22 were completed once more using the tablet. Subjects under the age of 6 years completed the MFM-20 in the usual way and items 18 and 22 once more using the tablet. The order (paper or tablet first) was determined using a computer-generated list of random numbers. Completions with the paper-based system and the tablet were performed with the same therapist. Scores of items 18, 19, and 22 obtained with the paper-based system were expressed as P-score, those obtained with the tablet T-score. An automatic scoring, expressed as A-score, was retrieved later on from the TabMe2 software.

An investigator meeting was organised before the start of the inclusions to ensure the correct completion and scoring of the MFM, as well as to provide training in the use of the TabMe2 software on the tablet. All the material was provided by the sponsor to each of the 5 study centres.

The day of MFM completion, demographic characteristics, diagnosis, as well as the score for each of the MFM items were collected. Therapists and subjects were also questioned about the relative of difficulty (easier, the same, or more difficult) of the item’s completion using the paper-based system and tablet.

### Statistical analysis

Quantitative variables were described by the number of subjects, the number of missing values, the median and the interquartile range [IQR]. Categorical variables were summarized by the number of each category and the number of missing values (missing values were not included in the denominator used for frequency computation).

With respect to the design of the study, as scores of items are ordinal [12], the assessment of agreement between the P-score and the T-score was evaluated using the weighted Cohen’s Kappa coefficient (quadratic weighting, so-called Fleiss-Cohen [13]) with its 95% confidence interval for each item. This coefficient express agreement as a number between 0 and 1 (0: no agreement; 1: perfect agreement).

The results were interpreted as suggested by Landis and Koch [14]: 0-0.20: slight, 0.21-0.40: fair, 0.41-0.60: moderate, 0.61-0.80: substantial, 0.81-1.00: almost perfect.

Exact and partial agreements for each item were displayed on a Bangdiwala chart [15]. This chart is a representation that displays agreement in paired categorical data where areas of various colour densities represent exact and partial agreements (respectively darker and lighter colours). The Bangdiwala chart reflects also a joint distribution of the scores, which gives a visual idea about the relative distributions of the scores of the two supports. The bias between these two supports is shown by the deviation of the rectangles from the 45° diagonal line within the larger square.

Analyses were conducted using SAS software version 9.3 for Windows (SAS Institute Inc., Cary, NC, USA).

## Results

### Characteristics of the population

Ninety-eight subjects (54 males and 44 females, aged between 5.0 and 60.0 years) were included in the study. The median age of the subjects was 16.2 years, and 55.1% were aged under 18 years of age. The 2 youngest subjects were less than 6 years old and completed an MFM-20; other subjects an MFM-32.

Duchenne or Becker muscular dystrophy were the most frequent (25.5% of subjects), followed by spinal muscular atrophy (16.3%) and myotonic dystrophy (15.3%). The median [IQR] total MFM score was 77.1% [53.1-85.4]; there were 32.7% of non-ambulant subjects and the median [IQR] MFM D1 score (standing and transfers) was 53.9% [12.8-74.4] (Table 1).

**Table 1.**
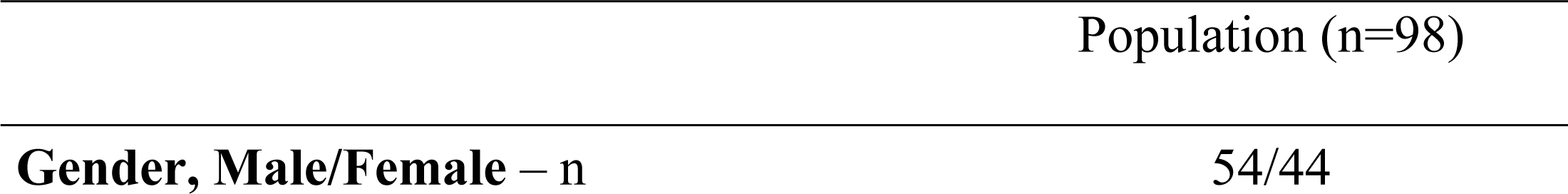
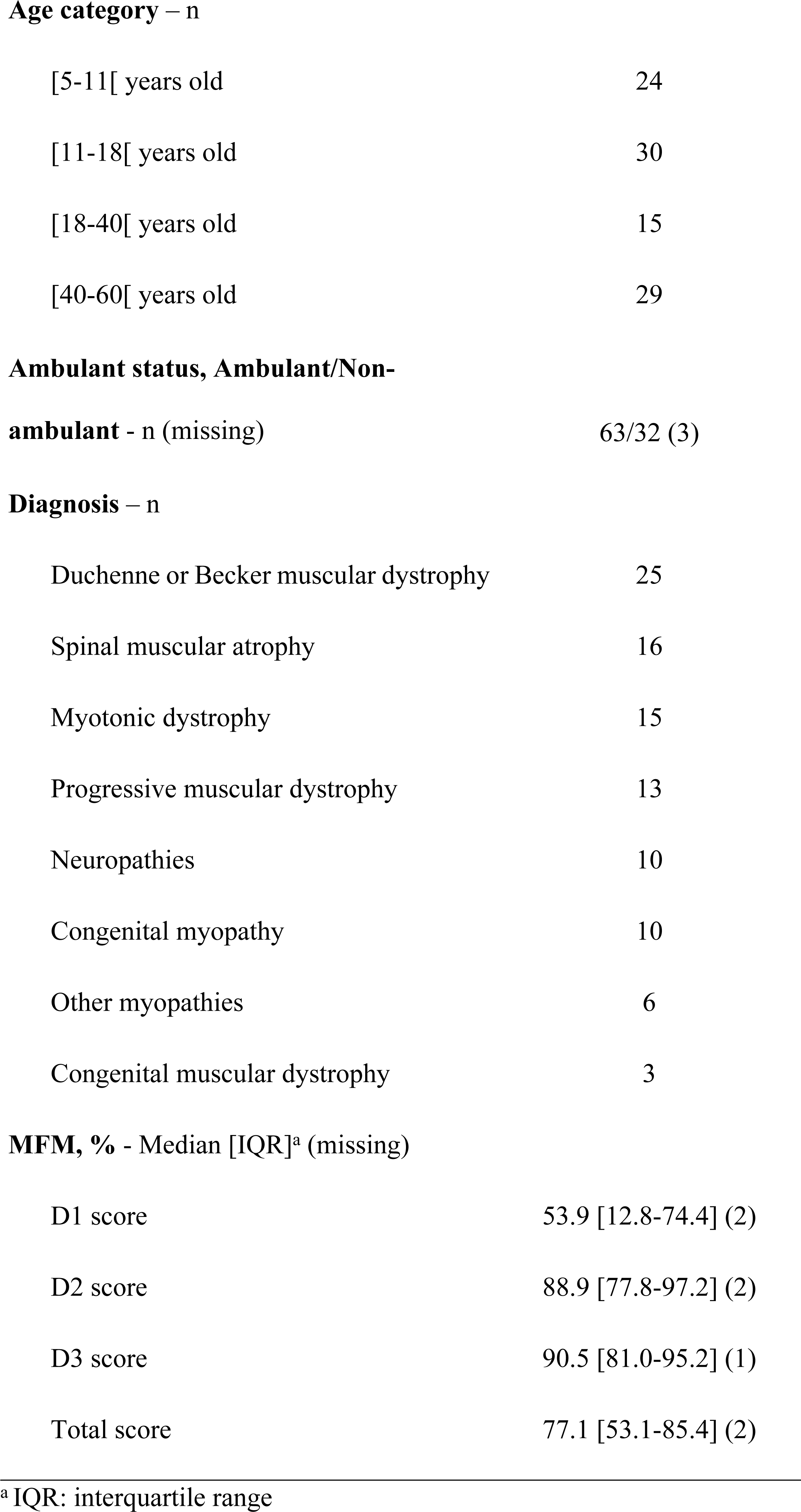
Subject characteristics.

### Agreements and disagreements between P and T-scores

Agreement between scores obtained with the paper-based support and the tablet (P-scores and T-scores, respectively) as assessed by the weighted Kappa coefficient was almost perfect for the scores of items 18 and 22 (*K*=0.93, and 0.95, respectively) and substantial for item 19 (*K*=0.70; Table 2).

**Table 2.** Percentage agreement and weighted Kappa coefficient between evaluation using the paper-based support or the tablet for item 18, 19 and 22.

|  | % agreement | Weighted Kappa coefficient | 95% confidence interval |
| --- | --- | --- | --- |
| Item 18 (n=96) | 93.7 | 0.93 | [0.86; 1.00] |
| Item 19 (n=96) | 74.0 | 0.70 | [0.55; 0.85] |
| Item 22 (n=98) | 98.0 | 0.95 | [0.88; 1.00] |

The agreement charts show the excellent agreement between the two modes of completion for items 18 and 22 (Figs 2A and 2C) and the good agreement for item 19, with a greater difficulty to complete this item on the tablet than on the paper-based support, which is reflected in the differences in the size of rectangles and the shift in relation to the 45° diagonal (Fig 2B).

**Fig 2.**
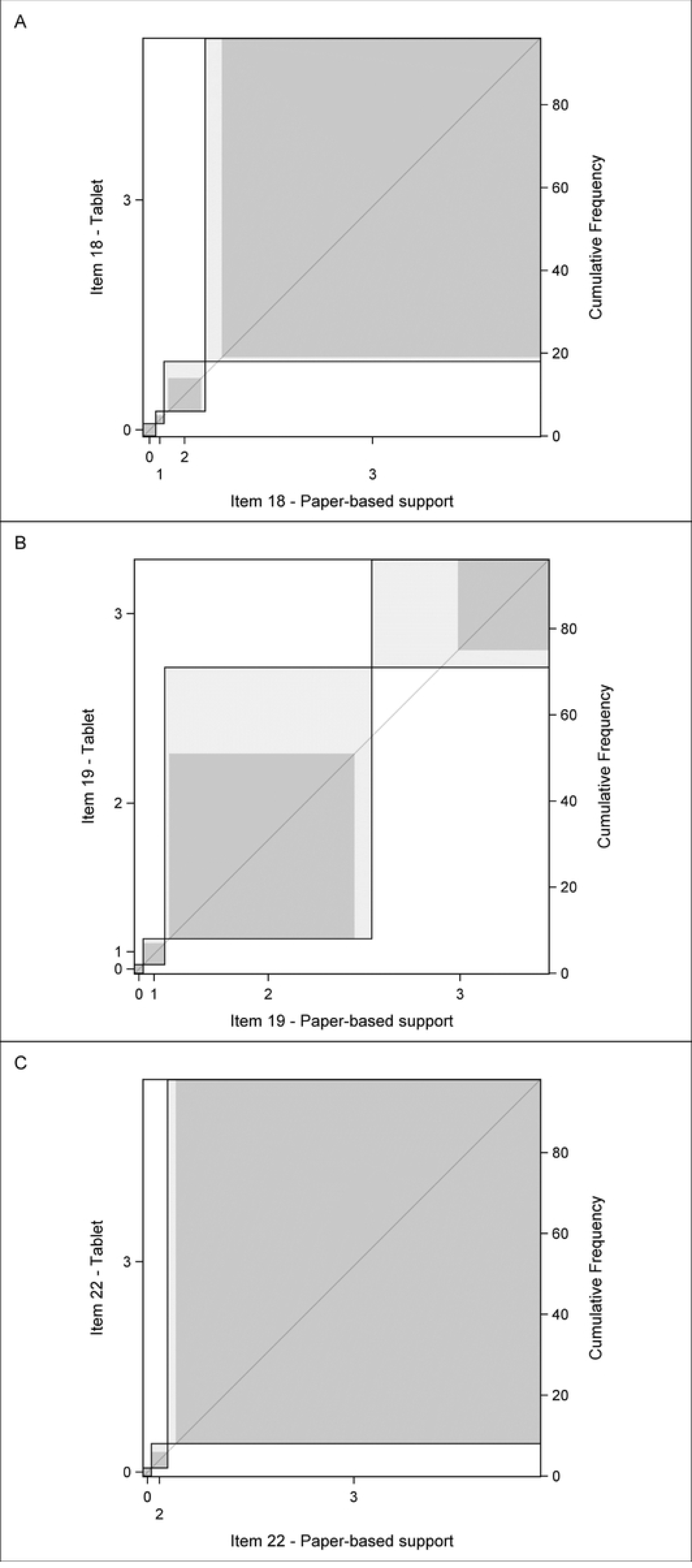
Bangdiwala’s agreement charts for item 18 (A), 19 (B) and 22 (C) comparing item scores between the two modes. Each item is represented by a distinct panel in which the four levels are represented by rectangles with one to two shades of grey. A deep grey area represents an exact agreement and a light grey area a partial agreement with a ‘one level away’ discrepancy. The bias between these two modes is shown by the deviation of the “rectangles” from the 45° diagonal line within the larger square.

The P-scores for items 18 and 22 were most frequently 3 (84.4% and 93.9% of cases, respectively); this was also the case for T-scores for the same items (81.2% and 91.8% of cases, respectively). For item 19 the most frequent P-score and T-score was 2 (50% and 65.6% of cases, respectively), followed by 3 (42.7% and 26.4% of cases, respectively (Table 3).

**Table 3.**
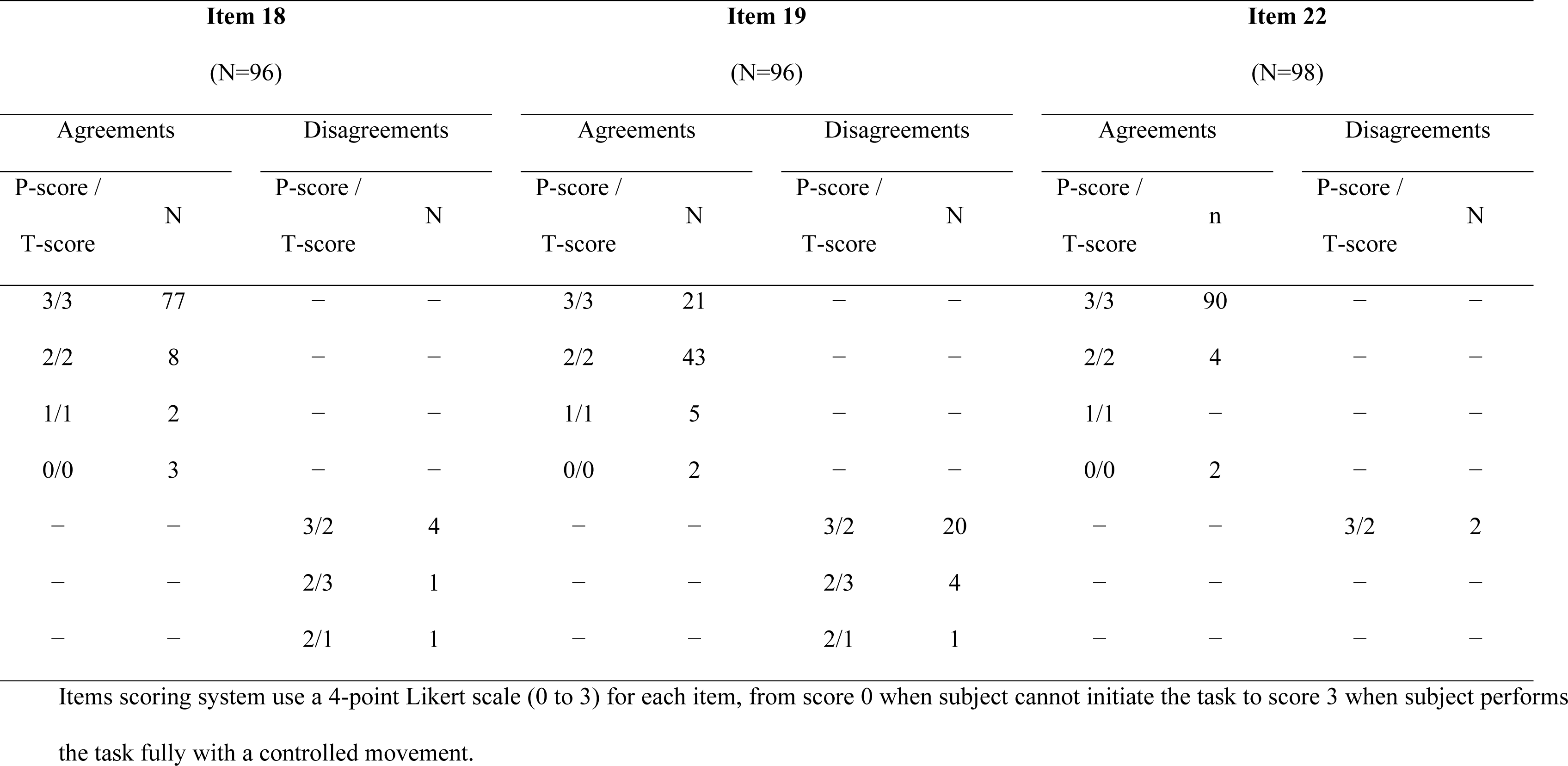
Agreements and disagreements between scores obtained during items 18, 19 and 22 completion using the Paper-based support (P-score) or the Tablet (T-score).

| Item 18 |  |  |  | Item 19 |  |  |  | Item 22 |  |  |  |
| --- | --- | --- | --- | --- | --- | --- | --- | --- | --- | --- | --- |
| (N=96) |  |  |  | (N=96) |  |  |  | (N=98) |  |  |  |
| Agreements |  | Disagreements |  | Agreements |  | Disagreements |  | Agreements |  | Disagreements |  |
| P-score / | N | P-score / | N | P-score / | N | P-score / | N | P-score / | n | P-score / | N |
| T-score |  | T-score |  | T-score |  | T-score |  | T-score |  | T-score |  |
| 3/3 | 77 | – | – | 3/3 | 21 | – | – | 3/3 | 90 | – | – |
| 2/2 | 8 | – | – | 2/2 | 43 | – | – | 2/2 | 4 | – | – |
| 1/1 | 2 | – | – | 1/1 | 5 | – | – | 1/1 | – | – | – |
| 0/0 | 3 | – | – | 0/0 | 2 | – | – | 0/0 | 2 | – | – |
| – | – | 3/2 | 4 | – | – | 3/2 | 20 | – | – | 3/2 | 2 |
| – | – | 2/3 | 1 | – | – | 2/3 | 4 | – | – | – | – |
| – | – | 2/1 | 1 | – | – | 2/1 | 1 | – | – | – | – |

Items scoring system use a 4-point Likert scale (0 to 3) for each item, from score 0 when subject cannot initiate the task to score 3 when subject performs the task fully with a controlled movement.

In all cases of disagreement, the difference was of one point. The most frequent disagreement concerned a P-score = 3 and a T-score = 2 for item 19 (20 disagreements). For items 18, 19, and 22, the score was identical in 71.4% of cases; the T-score was 1 point lower than the P-score in 24.5% of cases; the T score was 1 point higher than the P-score for 4.1% of cases (Table 3). For 1 subject, there was a difference in the scores of 2 items; the P-score for item 18 was 1 point higher than the T-score, and the T-score for item 19 was scored 1 point higher than the P-score.

### Level of difficulty as assessed by the subjects and the therapists for each item

For item 18, when the P and T-scores were identical, 44.6% of subjects found that the level of difficulty was identical (Fig 3A), and 70.5% of therapists did so (Fig 3B); 37.3% of subjects found that the task was less easy on the tablet (Fig 3A), as did 24.4% of therapists (Fig 3B).

**Fig 3.**
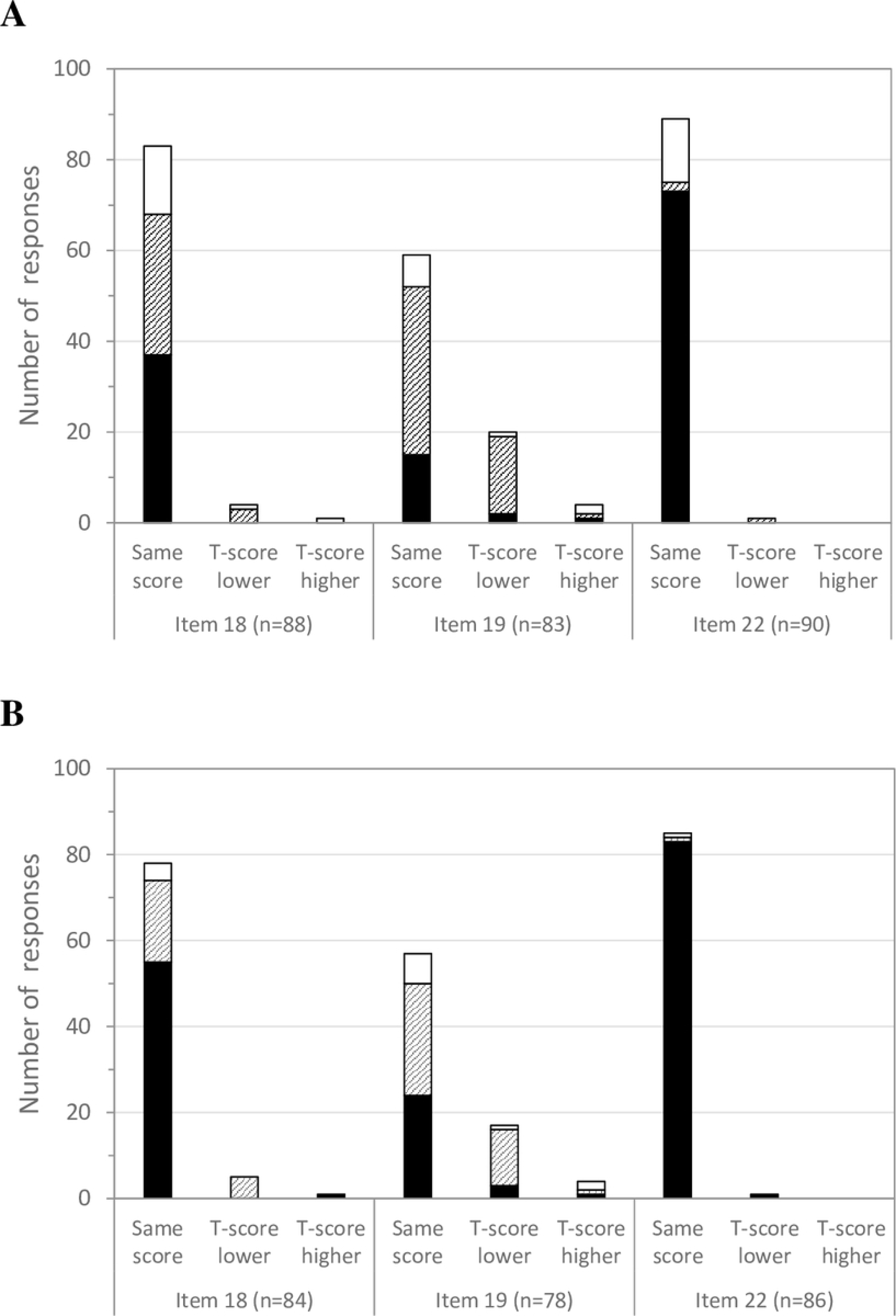
Assessment of the level of difficulty of the task from subjects (A) and therapists (B) when using the tablet compared to the paper-based support during the item 18, 19 and 22 completions. For each item, assessments are grouped according to each pair of scoring P-score and T-score results. Black bars indicate that the subjects or therapists considered that difficulty for completion of the task was the same level for the tablet and paper-based support, hatched bars indicate that they considered that it was less easy using the tablet, and white bars that it was easier using the tablet.

For item 19, when the P and T-scores were identical, 25.4% of subjects found that the level of difficulty was identical (Fig 3A), and 42.1% of therapists did so (Fig 3B); 62.7% of subjects found that it was less easy on the tablet (Fig 3A), and 45.6% therapists did so. When the T-score was lower, 85.0% found that it was less easy on the tablet (Fig 3A), as did 76.5% of therapists (Fig 3B).

For item 22, when the P and T-scores were identical, a large majority of subjects (82.0%) and therapists (97.6%) found that the level of difficulty was identical (Fig 3); 15.7% of subjects found that the level of difficulty were easier on the tablet (Fig 3A), as did 1.2% of therapists (Fig 3B).

When the P and T-scores were identical and the subjects found the task to be easier on the tablet subjects, 8 out of 14 subjects (57.1%) for item 18, 5 out of 7 subjects (71.4%) for item 19, and 10 out of 14 subjects (71.4%) for item 22 were under 11 years of age.

### Analysis of automatic score (A-score)

In order to assess the accuracy of TabMe2 automatic scoring procedures, we analysed from 23 subjects 29 completions of item 18, 28 of item 19, and 25 of item 22 (Table 4). Agreement between scores given by therapists when using the tablet (T-score) and the corresponding scores given automatically by the TabMe2 software based on the recorded tracks (A-score) were high for item 19 (89.3% of agreement) and item 22 (88.0% of agreement), respectively, and very low for item 18 (24.1% of agreement).

**Table 4.** Agreements and disagreements between scores of the therapist’s score on the tablet (T-score) and the automatic score (A-score) from the TabMe2 software.

| Item 18 |  |  |  | Item 19 |  |  |  | Item 22 |  |  |  |
| --- | --- | --- | --- | --- | --- | --- | --- | --- | --- | --- | --- |
| (N=29) |  |  |  | (N=28) |  |  |  | (N=25) |  |  |  |
| Agreements |  | Disagreements |  | Agreements |  | Disagreements |  | Agreements |  | Disagreements |  |
| T-score /<br>A-score | N | T-score /<br>A-score | N | T-score /<br>A-score | N | T-score /<br>A-score | N | T-score /<br>A-score | n | T-score /<br>A-score | N |
| 3/3 | 6 | — | — | 3/3 | 5 | — | — | 3/3 | 16 | — | — |
| 2/2 | 1 | — | — | 2/2 | 17 | — | — | 2/2 | 6 | — | — |
| 1/1 | — | — | — | 1/1 | 2 | — | — | 1/1 | — | — | — |
| 0/0 | — | — | — | 0/0 | 1 | — | — | 0/0 | — | — | — |
| — | — | 3/2 | 2 | — | — | 3/2 | 2 | — | — | 3/2 | 3 |
|  |  | 3/0 | 10 | — | — | — | — | — | — | — | — |
| — | — | 2/3 | 3 | — | — | — | — | — | — | — | — |
| — | — | — | — | — | — | 2/1 | 1 | — | — | — | — |
| — | — | 2/0 | 7 | — | — | — | — | — | — | — | — |

When we look at the reasons for the disagreements, there are calibration issues related to the margins of error accepted for all items (see S2 Table).

For item 18, specifically we often find issues with instructions given during the item completion. As outlined in the MFM manual, to obtain a score of 3 for item 18, the finger must go around the large CD, and not be on the edge of the CD. On tablet, participants often tried to follow precisely the mark representing the CD, resulting in traces unexpectedly entering the large circle, what had not been anticipated. For 6 cases an issue with the use of the digital interface occurred. Indeed, to help participants, we gave them the option of moving the CD around on the screen. However, when the CD was place to short from the edge of the tablet, pixel recording failures could occurred.

## Discussion

The present study provides evidence that supports the use of a tablet for the completion of items of the MFM dedicated to measure distal upper limb function. For the 3 studied items, the movement to be perform by the subject is the same for both modes of completion evaluated herein, and it involves the same motor compensations, making it possible to score and measure the motor task almost exactly in the same way. The strong agreements and the minimal differences in scores obtained in cases of disagreement allow the tablet to be used without a significant impact on subject follow-up. In case of differences in scoring, a maximum of one-point difference in scores when summing the scores of the 3 items was observed for each individual subjects, which is far beyond the minimal clinically important difference for the MFM that is reported to be between 2.4 and 5.9 points of the MFM total score [16–18]. This study with 98 patients is essential to authorise therapists and give them the confidence to use MFM in digital mode.

Although substantial, the weakest agreement between P-scores and T-scores was observed for item 19 (drawing loops in a predefined frame) and the item was frequently perceived as less easy when performed on a tablet, even when scores are identical. The majority difference in agreement, in terms of a 1-point reduction, is favourable to improving the sensitivity to changes in the item, which tends to have a ceiling effect. The subjects and therapists who found the task of item 19 on the tablet less easy often reported difficulties related to the stylus compared to the pencil: the stylus glided more or was more sensitive in 16 subjects and 3 therapists, a lack of experience using a stylus was reported by 8 subjects and 3 therapists, and a shaking problem when using the stylus was reported by 5 subjects and 5 therapists (*data not shown*). In a smaller proportion than for item 19, but still notable, subjects and therapists found the task of item 18 (going around the edge of a CD) less easy on the tablet. For these the absence of the rim created by the thickness of the CD was reported in 16 cases (*data not shown*), which could be used as a guide to turn around the CD for some subjects; however, despite the lack of a rim, agreement for this item was almost perfect. The third, item 22 (placing a finger on the drawings of the diagram), required no adaptation from subjects for the completion of the task. It was the item for which the highest agreement was found. Although a majority of score 3 was obtained whether using the tablet or paper-based support, around 15% of subjects found that it was easier on the tablet. A majority of these patients were under 11 years of age and 10 of these youngest subjects explained this by “it’s more interesting”, “better visibility”, “larger”, “the tablet is cool”, “the drawings look bigger”, or “I don’t know why but I prefer” (*data not shown*), which may be explained by better motivation and a task that was more enjoyable for them. Furthermore, the meaning of this item on paper-based support is not intuitive because it represents activity of daily living carried out on digital screens or keyboards. This item is more relevant to measure in digitally in the 21^st^ century, as is item 18.

The digitisation of the 3 items also paves the way for the automation of item scoring. This is all the more relevant given that items 18, 19 and 22 are part of the MFM items with the lower inter or intra-rater reliability[19]. The degree of agreements and the analysis of the disagreements between the T-score and A-score are promising for the development of very accurate automatic scoring. A score of 2 for item 18 can be given to a less well executed trace (which the algorithm could analyse), but also to subjects using the trunk or the whole of the upper limbs to compensate for limited hand motor skills. This type of compensation results in disagreements between T-Score and A-Score that wouldn’t be capture in the case of recording on a tablet at present.

Beyond an automatic scoring, the analyses of the traces on the tablet would be interesting to exploit in order to measure more finely the subjects’ hand skills [20]. While the presence of tremor is a hindrance in many activities of daily life and could lead to a reduction of scores from 3 to 2 for the 3 studied items, the characteristics of tremor are not currently assessed during the completion. Some teams have reported the possibility and the relevance of measuring it in other pathologies, such as Parkinson’s disease, multiple sclerosis, and essential tremor [21–24]. In the same way, a further computerized exploitation of the digital traces of recorded items could be interesting to quantify tremor in NMD, although this data has never been inventoried to date. The movement speed and accuracy could be assess as in the tablet-based application reported by Rabah et al. where most timing measures had excellent reliability in healthy participants or in the study of dexterity of Larsen et al. [25,26].

Lastly, the change of completion mode would be all the more significant in that these items were designed in 1998 to reproduce a function of use of digital peripherals (numeric keyboard, bank card terminal, touch screen, stylus, etc.). This would also make it possible to adapt to the disappearance of the marketing of the CDs necessary for the current manufacture of the test support.

Regarding the limitations of the study, the main one concerns the under-representation of 0 and 1 scores that precluded the analysis of the differences in scoring in such cases. This is likely to be due to the 3 studied items being among the easiest for subjects in MFM to complete. However, the population included in the present study reflects the targeted population for which the MFM is commonly used with all the variability of existing phenotypes, and only few very extremely weak patients were included. Another limitation is that this study concerns only an intra-rater reliability. Inter-rater reliability could also have been interesting to investigate as the tablet could change the way items are presented to the subject. However, the methodological choice was driven by reasons of feasibility, as it was easier to perform the tasks with both modes (tablet and paper) with the same therapist and by proceeding in this way the therapists were able to compare the completion.

## Conclusion

The switch from the original paper-based support to the tablet resulted in minimal and acceptable differences, and maintains a valid and reproducible measure of the 3 hand function items. We are moving forward to create a digital application called “MFM-play”, which will make it easier to facilitate patient follow-up, assist the therapist in the scoring process and integrate an interactive environment on the tablet to encourage children’s participation in MFMs. The realization of the items in digital mode also makes them compatible with solutions currently deployed to support mobile health systems for remote assessment.

## Data Availability

The de-identified research data supporting this publication could be obtained on request to the corresponding author

## Acknowledgments

The authors acknowledge Philip Robinson from DRS (Direction de la Recherche en Santé) of Hospices Civils de Lyon for help in manuscript preparation.

## Supporting information

**S1 File. Mathematical representation and processing carried out by TabMe2 software.**

**S2 Table. Illustrations of A-score for disagreements between scores of the therapist's score on the tablet (T-score) and the automatic score (A-score) from the TabMe2 software**

